# Mental health symptoms as preclinical indicators of dementia: a Whitehall II cohort study

**DOI:** 10.64898/2026.01.08.26343594

**Authors:** Monica Iyer, Aurore Fayosse, Mika Kivimaki, Gill Livingston, Archana Singh-Manoux, Charlotte Warren-Gash, Andrew Sommerlad, Séverine Sabia

**Affiliations:** London School of Hygiene and Tropical Medicine, Keppel Street, London WC1E 7HT, United Kingdom; Université Paris Cité and Université Sorbonne Paris Nord, Inserm U1153, INRAE, Centre for Research in Epidemiology and Statistics (CRESS), Epidemiology of Ageing and Neurodegenerative diseases (EpiAgeing), Paris, France; Division of Psychiatry, University College London, London, United Kingdom; North London NHS Foundation Trust, London, United Kingdom

## Abstract

**INTRODUCTION:** Changes in mental health symptoms, and their timing in the preclinical period of dementia, are not well characterised.

**METHODS:** We followed 5,495 Whitehall II participants (median age 68.5; 72.1% male) from their mental health symptoms assessment using the Clinical Interview Schedule–Revised (CIS-R) starting in 2012/13 to dementia diagnosis, death, or 2024. Linear mixed effects regression assessed CIS-R score changes preceding dementia. Flexible parametric models estimated associations of mental health symptoms with dementia.

**RESULTS:** Total CIS-R score increased (2.56 points [0.85-4.27]) in the 12 years preceding dementia. Having any mental health condition was associated with dementia in the short-term (HR at 3 years=4.04 [2.53-6.50]) but not the long-term (HR at 6 years=1.26 [0.63-2.49]). This pattern held for severe mental health conditions, concentration problems, depression, irritability, fatigue, anxiety, and worry.

**DISCUSSION:** Awareness of mental health symptoms as preclinical indicators of dementia in the short-term may support timely diagnosis of dementia.

## 1. INTRODUCTION

Timely dementia diagnosis is a priority to enable optimal management [1]. Dementia can be difficult to diagnose, as most diagnoses are made on a history of cognitive changes and functional impairment with neuroimaging and considering other potential causes of impairment without blood biomarkers [2]. Dementia has a long preclinical period, with changes in physical, cognitive, and neuropsychiatric health, along with changes in biomarkers. It is hypothesized that neuropsychiatric changes can serve as markers of preclinical dementia [3–5].

Much of the research which focuses on mental health conditions as prodromal or preclinical signs has focused on depression [6–10], though some studies have examined other mental health symptoms such as anxiety and personality changes [11–13]. One study found higher odds of dementia among those with prior depression, anxiety, and use of antipsychotics in the 10 years before dementia diagnosis [12], while another recent study identified six specific depressive symptoms as predictors of dementia risk over a 22-year follow-up period [14]. Meta-analyses considering other psychiatric disorders and risk of dementia reported mixed findings [15].

Many previous studies had follow-up of less than 5 years, which is insufficient to draw conclusions about whether changes in mental health cause or are due to dementia and, if the latter, their expected course over the long preclinical period [3,16,17]. They may be useful preclinical signs for dementia, which is necessary to understand the development of dementia and aid in early diagnosis. Knowledge of the timeline of changes in early symptoms of preclinical and prodromal dementia could help clinicians diagnose and care appropriately for people developing dementia.

We assessed the association between mental health conditions and subsequent dementia in the United Kingdom (UK) Whitehall II cohort study over a follow-up period of 12 years. The objectives were to: 1) Examine the change in overall mental health and specific mental health symptoms during up to 12 years preceding diagnosis of dementia and, 2) Examine the association of overall mental health and specific mental health symptoms with risk of dementia over time from mental health symptoms assessment.

## 2. METHODS

### 2.1. Study population

The Whitehall II study cohort consists of 10,308 participants recruited from the British Civil Service in 1985, who were aged 35-55 at baseline [17]. The study consists of a self-administered questionnaire assessing demographic, social, lifestyle, and clinical variables, and a clinical screening for various health measures, carried out every 4-5 years after baseline. Written informed consent from participants and research ethics approvals were renewed at each contact. Ethics approval is from the University College London Hospital Committee on the Ethics of Human Research, reference number 85/0938.

### 2.2. Mental health variables

Our study population consisted of all participants, who completed the self-administered computerized version of the Clinical Interview Schedule-Revised (CIS-R) to measure mental health and were free of dementia at the 2012-13 clinical visit [18]. Participants completed the CIS-R, a structured diagnostic interview tool validated in general populations to capture mental health conditions, at the 2012-13 and 2015-16 clinical visits. It has a sensitivity and specificity for any mental health disorder, with a cutoff score of ≥12 of 74% and 98%, respectively [19] and also generates 14 different mental health symptom subscores with ranges from 0-4 (except depressive ideas, which is 0-5), so the total score ranges from 0-57. The 14 symptoms are: anxiety, compulsions, concentration problems, depression, depressive ideas, fatigue, irritability, obsessions, panic, phobias, sleep problems, somatic symptoms, worry, and worry over physical health.

### 2.3. Dementia diagnosis

Dementia diagnostic status was ascertained from three linked electronic health record sources: hospital-recorded dementia from Hospital Episode Statistics (HES) data, dementia diagnosed in secondary mental healthcare services through the Mental Health Services Dataset (MHSD), and mortality data with dementia as a cause of death. The algorithm for defining dementia is in Supplemental Table 1. Dementia diagnoses were available until March 31, 2024. HES dementia data up to 2016 had sensitivity of 78% and specificity of 92% [20]; our data is likely to be more accurate given the addition of the MHSD and mortality data and trends of increasing electronic healthcare record accuracy [21].

### 2.4. Covariates

Covariates were extracted from the 2012-13 wave that included a questionnaire and clinical examination, and from linkage to electronic records; they included demographic, lifestyle, and health-related factors. The demographic variables were age, sex, ethnicity (White, non-White), marital status (Married/cohabitating or not), and education level (None or O level, A level, BA/BSc or more). Lifestyle variables include self-reported fruit and vegetable intake, physical activity, alcohol consumption, and smoking. Health related factors included body mass index (BMI) measured at the clinic, and diagnoses of diabetes, hypertension, or multimorbidity, assessed through a combination of questionnaire, HES data, previous Whitehall II screening results, mental health records, mortality records, or cancer registries. Multimorbidity was defined as having diagnoses of two or more chronic conditions among: cancer, coronary heart disease, chronic kidney disease, chronic obstructive pulmonary disease, heart failure, liver disease, Parkinson’s, rheumatoid arthritis, stroke, or osteoarthritis. The criteria and data sources used for each of the conditions are in Supplemental Table 1. Some participants had missing covariate information (up to 3.7% of 2012-13 clinical visit participants), in which case participant’s information from previous visits was used if available.

### 2.5. Statistical Analysis

We described the demographic, lifestyle, and health-related characteristics of the study population, overall and stratified by baseline mental health status and whether participants had a dementia diagnosis in follow-up, with differences in groups assessed using either Chi² or t-test, as appropriate. Participants were followed from the date of their CIS-R completion in 2012-13 until dementia diagnosis, death, or the end of the study period (March 2024), whichever came first. We used the CIS-R overall score and the symptom subscores as continuous variables for Objective 1. For Objective 2, we used binary variables to indicate any mental health condition (overall score of ≥12), severe mental health condition (overall score of ≥ 18) and symptoms of each mental health subscore (symptom subscore of ≥2). Due to the low number of participants with subscores of ≥2, we excluded panic and phobias from the statistical analyses.

#### 2.5.1. Objective 1. Change in mental health in the years preceding dementia

To estimate the change in mental health disorders (overall and by subscores) in the years before dementia, we used linear mixed effect models [22]. We used linear regression with CIS-R total score as the dependent variable at 2012-13 and 2015-16 visits, and time to dementia, age at dementia diagnosis, sex, and ethnicity as the independent variables with a random effects term on the individuals. We tested whether the slope of the change in CIS-R score over time changed at different time points through the introduction of a breakpoint in the linear mixed effect model [23]. To compare the change in CIS-R score among those with and without dementia, we matched individuals with dementia to controls. Each dementia case was matched based on age (±2 years) and sex with up to five controls who were dementia free at the dementia diagnosis date of the corresponding case. We then ran the linear mixed effect model on CIS-R score with fixed effects for time, dementia status, and their interaction, sex, age, and ethnicity and random intercepts for individuals. The interaction between time and dementia status indicated whether rates of change in CIS-R score differed between cases and controls. We repeated this analysis for each individual subscore which showed evidence of change among cases in the initial analysis, and used the time identified as meaningful in the breakpoint analysis.

#### 2.5.2. Objective 2. Association between mental health symptoms and dementia

We initially ran a Cox proportional hazards model and found that proportional hazards assumption was not met through inspection of the Schoenfeld residuals plot. We therefore used flexible parametric models, to allow flexibility in the proportional hazards assumption, to estimate and plot the hazard ratio of mental health symptoms (separately for any mental health condition, severe mental health condition, and each subscore) and risk of subsequent dementia over time. In the flexible parametric model, we used time since the measure of the CIS-R at the 2012-13 visit as the timescale and adjusted the models for demographic, lifestyle, and clinical variables. We extracted the hazard ratio at each year of follow-up for dementia associated with the presence of any mental health condition in 2012-13. For the individual mental health symptoms, we plot the hazard ratio over 12 years and extracted the hazard ratio for dementia at 3 years of follow-up in the main text to provide an interpretable and clinically meaningful estimate of risk. We did a complete-case analysis so excluded seven participants missing ethnicity and one missing BMI information.

#### 2.5.3. Sensitivity analyses

Of the Whitehall II study participants who were alive and without dementia at the time of the 2012-13 visit, 84.1% were included in our analyses. We conducted a sensitivity analysis using Inverse Probability Weighting (IPW) to account for missing data [23,24]. We used Cox regression to calculate a hazard ratio for any mental health condition and dementia in the original unweighted population and compared it with the hazard ratio of the IPW population. A description of the calculation of the weights is provided in Supplementary Methods.

## 3. RESULTS

### 3.1. Descriptive Results

The 5,495 participants who were free of dementia and completed the CIS-R at the 2012-13 visit were included (flow chart in Supplementary Figure 1). They were followed for a median 11.25-years (Interquartile range [IQR]: 10.68, 11.63). Table 1 presents the baseline characteristics of the study population, overall and stratified by presence of any mental health condition at the 2012-13 visit and by incident dementia over the follow-up. The median age at the 2012-13 visit was 68.5 years (IQR: 64.8, 74.2), and the majority of participants were male (72.1%) and White (92.7%) (Table 1). At baseline, 454 (8.3%) people had any mental health condition (total CIS-R score ≥12), and they were more likely to be female, non-white, not married, and generally less healthy compared to those without a mental health condition at baseline (Table 1). During follow-up, 455 (8.3%) people developed dementia (Table 1), with the median (IQR) time to dementia being 7.15 (4.43 to 9.47) years. People who developed dementia were on average older and less healthy than those who did not develop dementia during the follow-up period (Table 1).

**Table 1.** Baseline characteristics of study population, overall and by mental health condition and dementia status.

| Characteristic | Overall<br>N=5,495 | No mental health<br>condition<br>N=5,041 | Mental health<br>condition (total<br>CIS-R score $\geq 12$)<br>N=454 | p-value<sup>1</sup> | No dementia<br>N=5,040 | Dementia<br>N=455 | p-value<sup>1</sup> |
| --- | --- | --- | --- | --- | --- | --- | --- |
| Age, median (IQR) | 68.5 (64.8, 74.2) | 68.5 (64.8, 74.2) | 68.4 (64.7, 73.7) | 0.5 | 67.9 (64.5, 73.4) | 75.6 (70.3, 78.7) | <0.001 |
| Sex, Male n (%) | 3,964 (72.1%) | 3,704 (73.5%) | 260 (57.3%) | <0.001 | 3,650 (72.4%) | 314 (69.0%) | 0.130 |
| Ethnicity, White n (%) | 5,096 (92.7%) | 4,691 (93.1%) | 405 (89.2%) | 0.004 | 4,686 (93.0%) | 410 (90.1%) | 0.023 |
| Marital status,<br>Married/cohabitating n (%) | 4,081 (74.3%) | 3,800 (75.4%) | 281 (61.9%) | <0.001 | 3,768 (74.8%) | 313 (68.8%) | 0.006 |
| Education, n (%) |  |  |  | 0.6 |  |  | <0.001 |
| O level or none | 2,285 (41.6%) | 2,088 (41.4%) | 297 (65.4%) |  | 2,046 (40.6%) | 239 (52.5%) |  |
| A level | 1,492 (27.2%) | 2,369 (47.0%) | 123 (27.1%) |  | 1,395 (27.7%) | 97 (21.3%) |  |
| BA/BSc or more | 1,718 (31.3%) | 1,584 (31.4%) | 134 (29.5%) |  | 1,599 (31.7%) | 119 (26.2%) |  |
| Fruit and vegetable intake,<br>n (%) |  |  |  | 0.013 |  |  | <0.001 |
| Daily or less | 2,317 (42.2%) | 2,100 (41.7%) | 217 (47.8%) |  | 2,079 (41.3%) | 238 (52.3%) |  |
| More than daily | 3,178 (57.8%) | 2,941 (58.3%) | 237 (52.2%) |  | 2,961 (58.8%) | 217 (47.7%) |  |
| Physical activity (hours per<br>week), n (%) |  |  |  | <0.001 |  |  | <0.001 |
| 0 | 476 (8.7%) | 406 (8.1%) | 70 (15.4%) |  | 419 (8.3%) | 57 (12.5%) |  |
| <2.5 | 1,931 (35.1%) | 1,753 (34.8%) | 178 (39.2%) |  | 1,753 (34.8%) | 178 (39.1%) |  |
| $\geq 2.5$ | 3,088 (56.2%) | 2,882 (57.2%) | 206 (45.4%) | | 2,868 (56.9%) | 220 (48.4%) | |
| Alcohol units per week, n<br>(%) |  |  |  | <0.001 |  |  | 0.10 |
| 0 | 1,149 (20.9%) | 1,055 (20.9%) | 144 (31.7%) |  | 1,039 (20.6%) | 110 (24.2%) |  |
| 1-14 | 3,074 (55.9%) | 2,865 (56.8%) | 209 (46.0%) |  | 2,820 (56.0%) | 254 (55.8%) |  |
| >14 | 1,272 (23.1%) | 1,171 (23.2%) | 101 (22.2%) |  | 1,181 (23.4%) | 91 (20.0%) |  |
| Smoking, n (%) |  |  |  | 0.079 |  |  | 0.70 |
| Never | 2,658 (48.4%) | 2,454 (48.7%) | 204 (44.9%) |  | 2,439 (48.4%) | 219 (48.1%) |  |
| Ex-smoker | 2,645 (48.1%) | 2,418 (48.0%) | 227 (50.0%) |  | 2,422 (48.1%) | 223 (49.0%) |  |
| Current smoker | 192 (3.5%) | 169 (3.4%) | 23 (5.1%) |  | 179 (3.6%) | 13 (2.9%) |  |
| BMI (kg/m<sup>2</sup>), n (%) |  |  |  | 0.2 |  |  | 0.20 |
| <25 | 2,073 (37.7%) | 1,911 (37.9%) | 162 (35.7%) |  | 1,887 (37.4%) | 186 (40.9%) |  |
| 25-<30 | 2,348 (42.7%) | 2,159 (42.8%) | 189 (41.6%) |  | 2,155 (42.8%) | 193 (42.4%) |  |
| ≥30 | 1,073 (19.5%) | 970 (19.2%) | 103 (22.7%) |  | 997 (19.8%) | 76 (16.7%) |  |
| Diabetes, n (%) | 730 (13.3%) | 655 (13.0%) | 75 (16.5%) | 0.041 | 651 (12.9%) | 79 (17.4%) | 0.007 |
| Hypertension, n (%) | 3,443 (62.7%) | 3,160 (62.7%) | 283 (62.3%) | >0.9 | 3,102 (61.5%) | 341 (74.9%) | <0.001 |
| Multimorbidity, n (%) | 416 (7.6%) | 366 (7.3%) | 50 (11.0%) | 0.005 | 371 (7.4%) | 45 (9.9%) | 0.051 |

BMI = Body Mass Index; CIS-R = Clinical Interview Schedule, Revised; IQR = Interquartile Range
<sup>1</sup>p-value: Chi squared test for association for categorical variables and t-test for continuous variables
Note: Seven participants were missing ethnicity; one was missing BMI. They were excluded from the main analysis.

The median total CIS-R score at the 2012-13 visit was 2 (IQR: 0, 5), and 183 (3.3%) had a severe mental health condition (total score ≥18). The most common symptom was sleep problems, followed by fatigue, irritability, and worry (Table 2). Table 2 also shows the prevalence of mental health symptoms in 2012-13 according to dementia incidence by the end of follow-up. Of the 455 participants with dementia between 2012-13 and the end of follow-up, 314 developed dementia between their participation in the CIS-R at the 2015-16 visit and the end of follow-up (Supplementary Table 2).

**Table 2.**
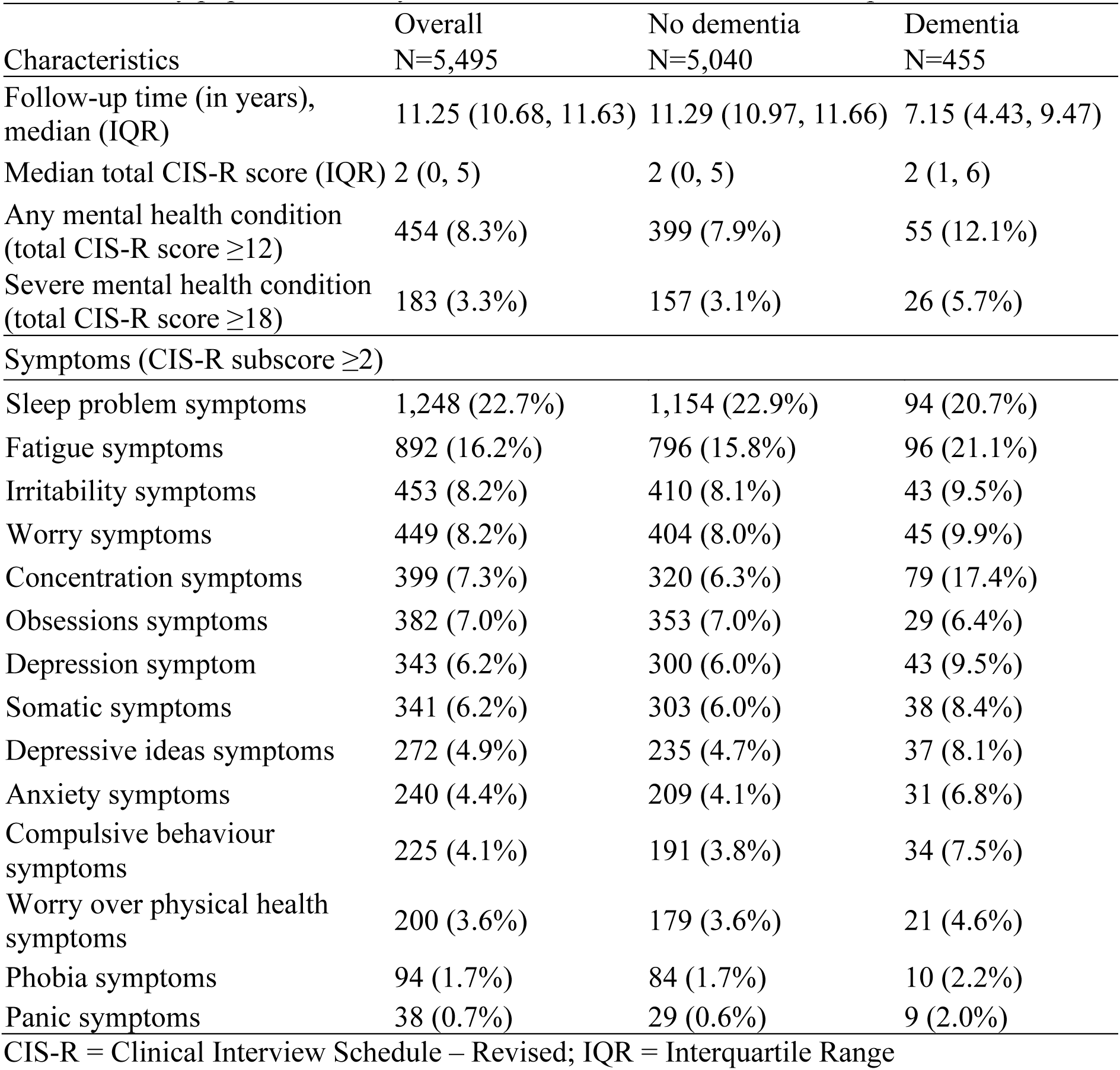
Prevalence of mental health symptoms (overall and each subscore) at 2012-13 visit, in the full study population and by dementia status at the end of follow-up.

| Characteristics | Overall<br>N=5,495 | No dementia<br>N=5,040 | Dementia<br>N=455 |
| --- | --- | --- | --- |
| Follow-up time (in years), median (IQR) | 11.25 (10.68, 11.63) | 11.29 (10.97, 11.66) | 7.15 (4.43, 9.47) |
| Median total CIS-R score (IQR) | 2 (0, 5) | 2 (0, 5) | 2 (1, 6) |
| Any mental health condition (total CIS-R score $\geq 12$) | 454 (8.3%) | 399 (7.9%) | 55 (12.1%) |
| Severe mental health condition (total CIS-R score $\geq 18$) | 183 (3.3%) | 157 (3.1%) | 26 (5.7%) |
| Symptoms (CIS-R subscore $\geq 2$) | | | |
| Sleep problem symptoms | 1,248 (22.7%) | 1,154 (22.9%) | 94 (20.7%) |
| Fatigue symptoms | 892 (16.2%) | 796 (15.8%) | 96 (21.1%) |
| Irritability symptoms | 453 (8.2%) | 410 (8.1%) | 43 (9.5%) |
| Worry symptoms | 449 (8.2%) | 404 (8.0%) | 45 (9.9%) |
| Concentration symptoms | 399 (7.3%) | 320 (6.3%) | 79 (17.4%) |
| Obsessions symptoms | 382 (7.0%) | 353 (7.0%) | 29 (6.4%) |
| Depression symptom | 343 (6.2%) | 300 (6.0%) | 43 (9.5%) |
| Somatic symptoms | 341 (6.2%) | 303 (6.0%) | 38 (8.4%) |
| Depressive ideas symptoms | 272 (4.9%) | 235 (4.7%) | 37 (8.1%) |
| Anxiety symptoms | 240 (4.4%) | 209 (4.1%) | 31 (6.8%) |
| Compulsive behaviour symptoms | 225 (4.1%) | 191 (3.8%) | 34 (7.5%) |
| Worry over physical health symptoms | 200 (3.6%) | 179 (3.6%) | 21 (4.6%) |
| Phobia symptoms | 94 (1.7%) | 84 (1.7%) | 10 (2.2%) |
| Panic symptoms | 38 (0.7%) | 29 (0.6%) | 9 (2.0%) |

CIS-R = Clinical Interview Schedule – Revised; IQR = Interquartile Range

### 3.2. Objective 1. Change in mental health in the years preceding dementia

Using the CIS-R total scores from the 2012-13 and 2015-16 visits, among those who developed dementia during the follow-up, the average estimated increase in CIS-R score per 12 years adjusted for age at dementia diagnosis, sex, and ethnicity was 2.56 (95% confidence interval [CI]: 0.85, 4.27; p-value: 0.003). We tested whether the slope changed at different breakpoints and found evidence that the slope increased 6 years before dementia diagnosis (p-value for a change in slope: 0.042). When we added a breakpoint to the linear mixed effect model, there was no evidence of a change in overall CIS-R score from years 12 to 6 years before diagnosis (6-year change in score: 0.168, 95% CI: −1.20, 1.54, p-value: 0.81), but there was evidence of a change in the score in the 6 years preceding a dementia diagnosis (6-year change: 2.61, 95% CI: 1.08, 4.13, p-value: <0.001). This 6-year change was greater than the 6-year change of the matched controls, for whom there was no evidence of an increase in score (6-year change for controls: 0.27, 95% CI: −0.13, 0.68, p-value for difference between cases and controls: <0.001).

Six symptoms showed increases in CIS-R subscore in the 12 years preceding a dementia diagnosis: concentration problems, fatigue, depression, irritability, depressive ideas, and worry over physical health. When we added a breakpoint at 6 years before dementia diagnosis, none of the symptoms had evidence of a change in CIS-R subscore from 12 to 6 years preceding dementia diagnosis, but seven symptoms had evidence of an increase in CIS-R subscore in the 6 years preceding a dementia diagnosis: concentration problems, fatigue, depression, irritability, depressive ideas, worry over physical health, and anxiety. However, there was weak to no evidence of there being a change in slope from the pre to post breakpoint, apart from the concentration problems symptom for which there was evidence of a change in slope (p-value: 0.013). The change in CIS-R score in the 6 years before dementia diagnosis for the seven symptoms were all greater than the change in score for the matched controls, though there was less of a difference between cases and controls for depression and depressive ideas (p-value for depression: 0.06; depressive ideas: 0.05; irritability: 0.02; fatigue, concentration, anxiety, and worry over physical health: <0.001) (Table 3).

**Table 3.**
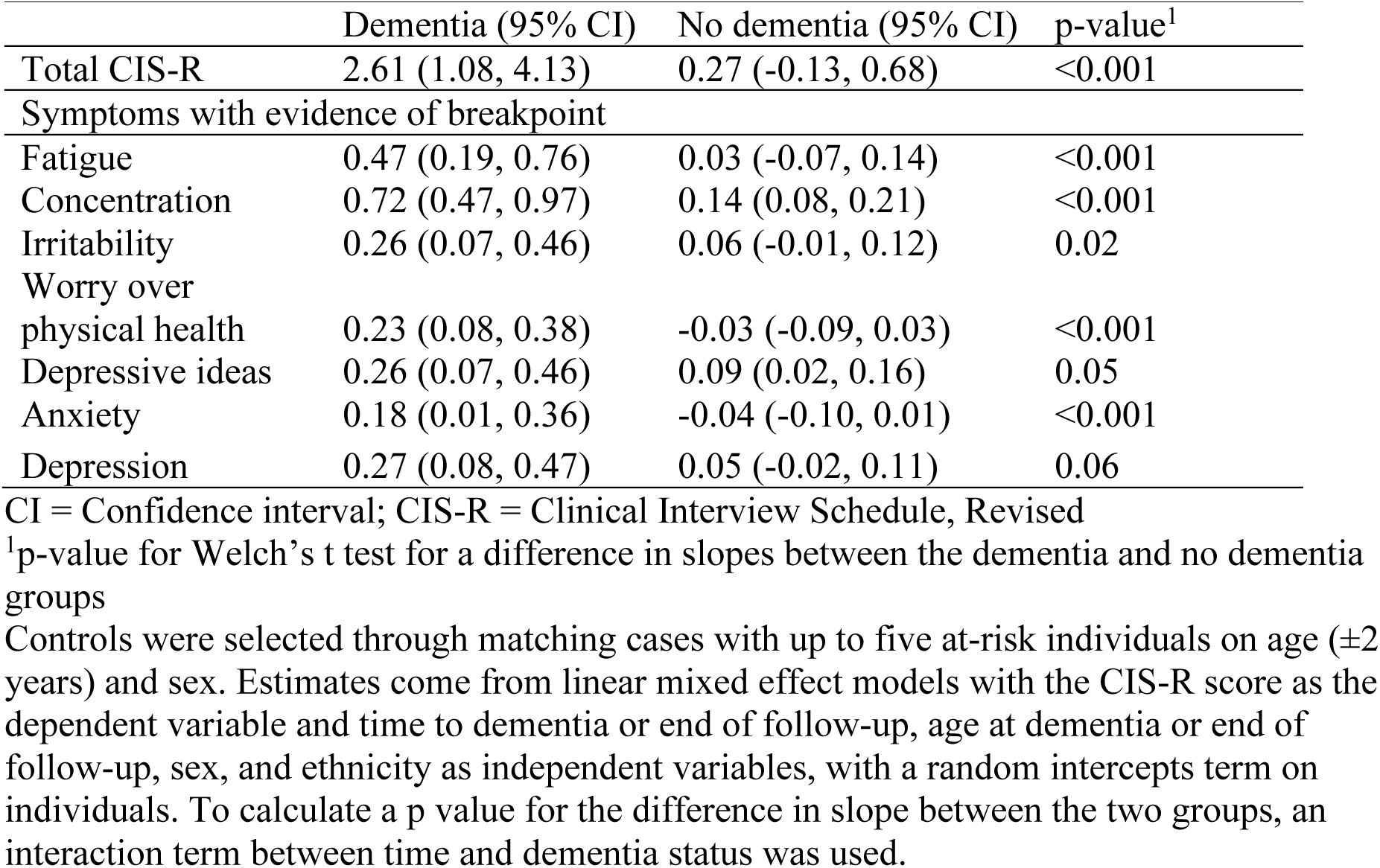
Change in CIS-R score in 6 years before dementia diagnosis (dementia group) or before end of follow-up or death (no dementia group)

**Table 4.**
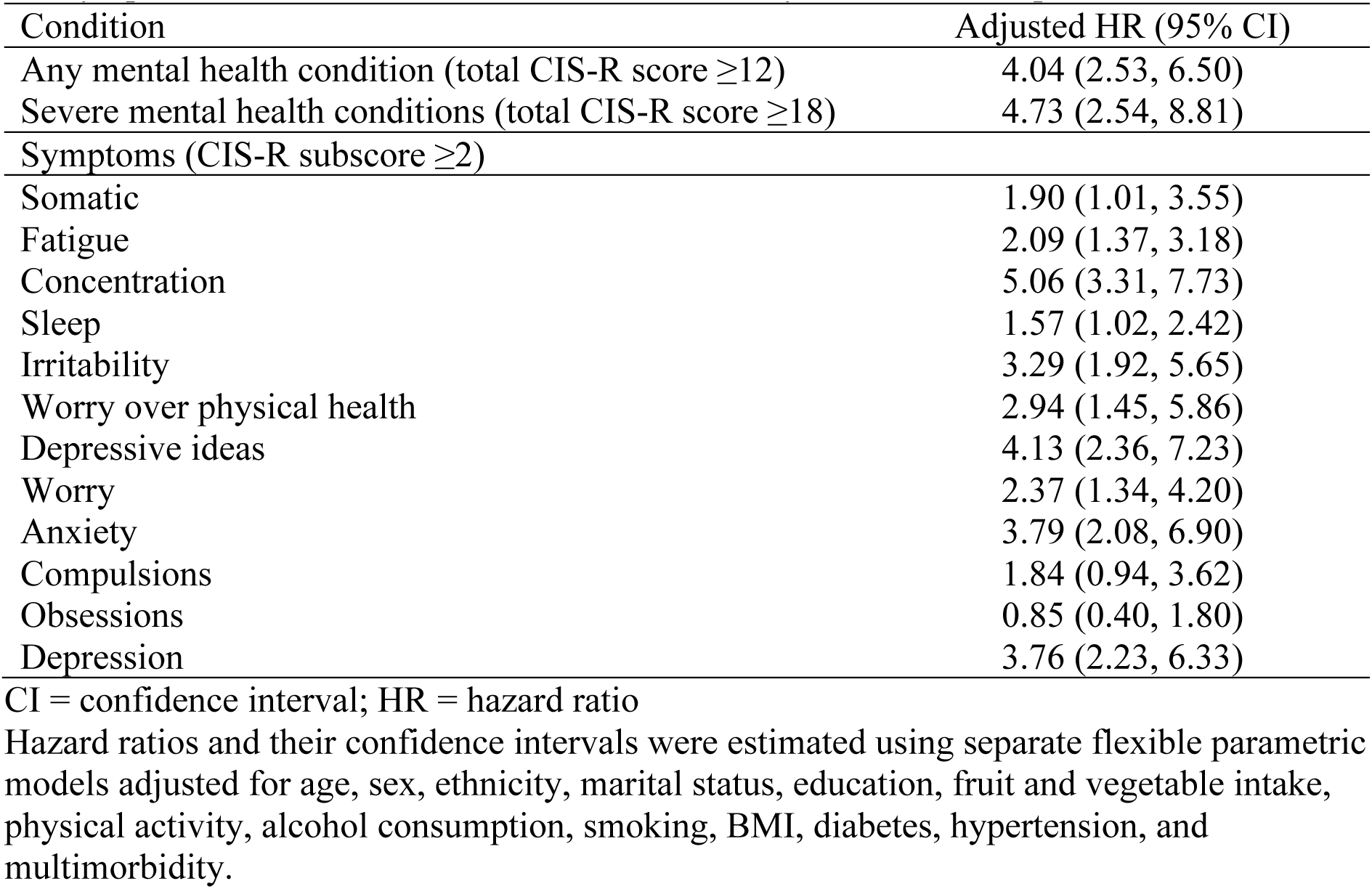
Association between the any mental health condition, severe mental health condition and symptom subscores and risk of dementia at three years of follow-up.

| Condition | Adjusted HR (95% CI) |
| --- | --- |
| Any mental health condition (total CIS-R score $\geq 12$) | 4.04 (2.53, 6.50) |
| Severe mental health conditions (total CIS-R score $\geq 18$) | 4.73 (2.54, 8.81) |
| Symptoms (CIS-R subscore $\geq 2$) | |
| Somatic | 1.90 (1.01, 3.55) |
| Fatigue | 2.09 (1.37, 3.18) |
| Concentration | 5.06 (3.31, 7.73) |
| Sleep | 1.57 (1.02, 2.42) |
| Irritability | 3.29 (1.92, 5.65) |
| Worry over physical health | 2.94 (1.45, 5.86) |
| Depressive ideas | 4.13 (2.36, 7.23) |
| Worry | 2.37 (1.34, 4.20) |
| Anxiety | 3.79 (2.08, 6.90) |
| Compulsions | 1.84 (0.94, 3.62) |
| Obsessions | 0.85 (0.40, 1.80) |
| Depression | 3.76 (2.23, 6.33) |

CI = confidence interval; HR = hazard ratio
Hazard ratios and their confidence intervals were estimated using separate flexible parametric models adjusted for age, sex, ethnicity, marital status, education, fruit and vegetable intake, physical activity, alcohol consumption, smoking, BMI, diabetes, hypertension, and multimorbidity.

Figure 1A illustrates the change in CIS-R subscore before and after the breakpoint for these symptoms and the β estimate with 95% CIs for the post-breakpoint segment (the 6 years preceding dementia diagnosis). Figure 1B illustrates the trend line of the symptoms which had no evidence of a change in CIS-R score before or after the breakpoint: compulsions, obsessions, sleep problems, somatic, and worry. Due to there being no evidence of a breakpoint, we report the overall trend line with a β estimate and corresponding 95% CI for a 12-year change (the full follow-up period).

**Figure 1.**
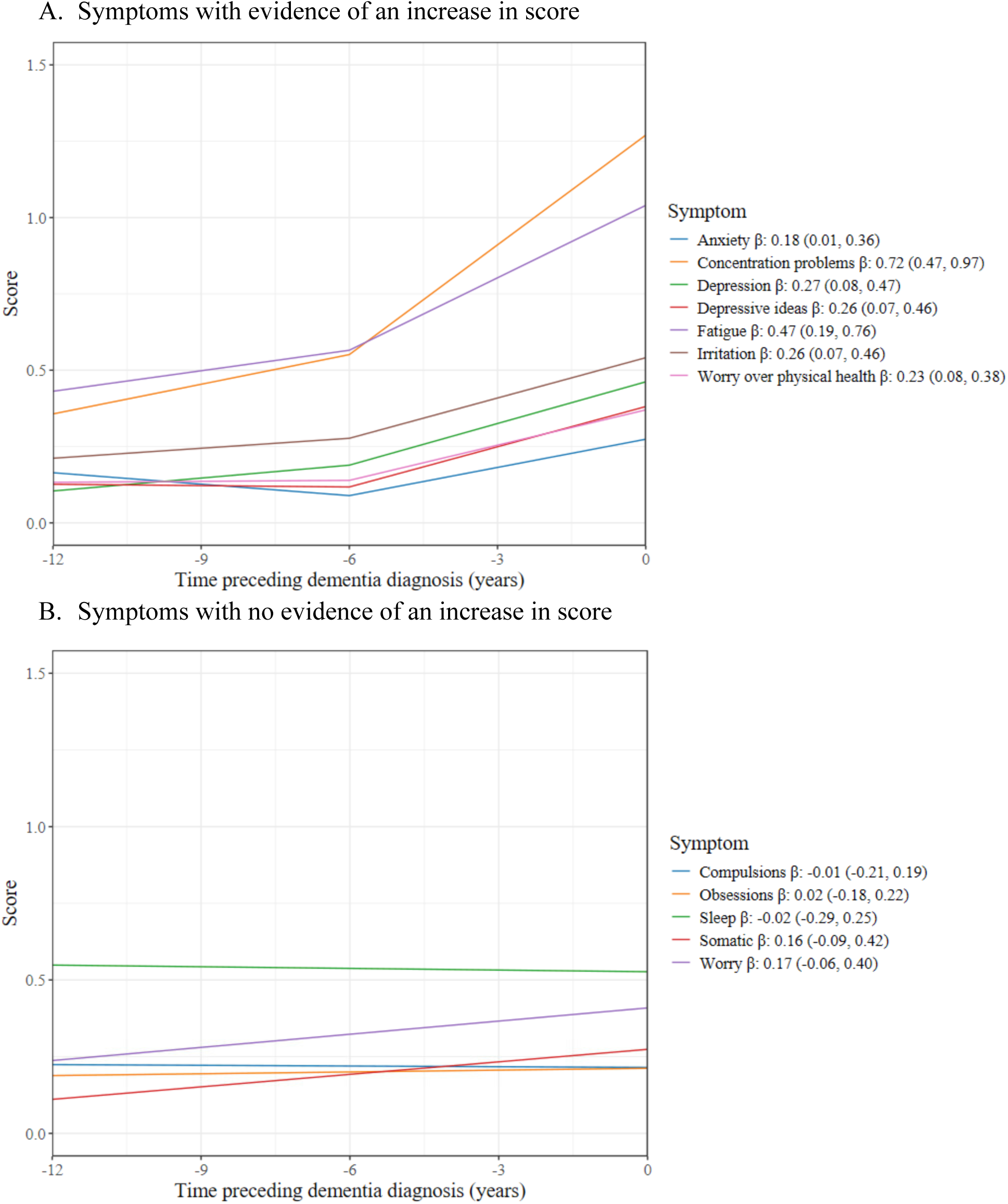
Change in CIS-R symptom subscore in years preceding dementia β: Average change in CIS-R score in 6 years preceding dementia diagnosis (95% CI) in Panel A, and average change in CIS-R score in 12 years preceding dementia diagnosis (95% CI) in Panel B Estimates come from linear mixed effect models with the mental health score as the dependent variable and time to dementia diagnosis, age at dementia diagnosis, sex, and ethnicity as independent variables. Estimates correspond to predicted values for the referent categories of a White man aged 81.2 years at dementia diagnosis (mean age at dementia in the sample).

### 3.3. Objective 2. Association between mental health symptoms and dementia

Figure 2 displays the hazard ratios (and 95% CIs) for dementia incidence over the time since the measure of the CIS-R in 2012-13 associated with any mental health condition, severe mental health condition (Panel A), and each symptom (Panel B), adjusted for demographic, lifestyle, and clinical factors.

**Figure 2.**
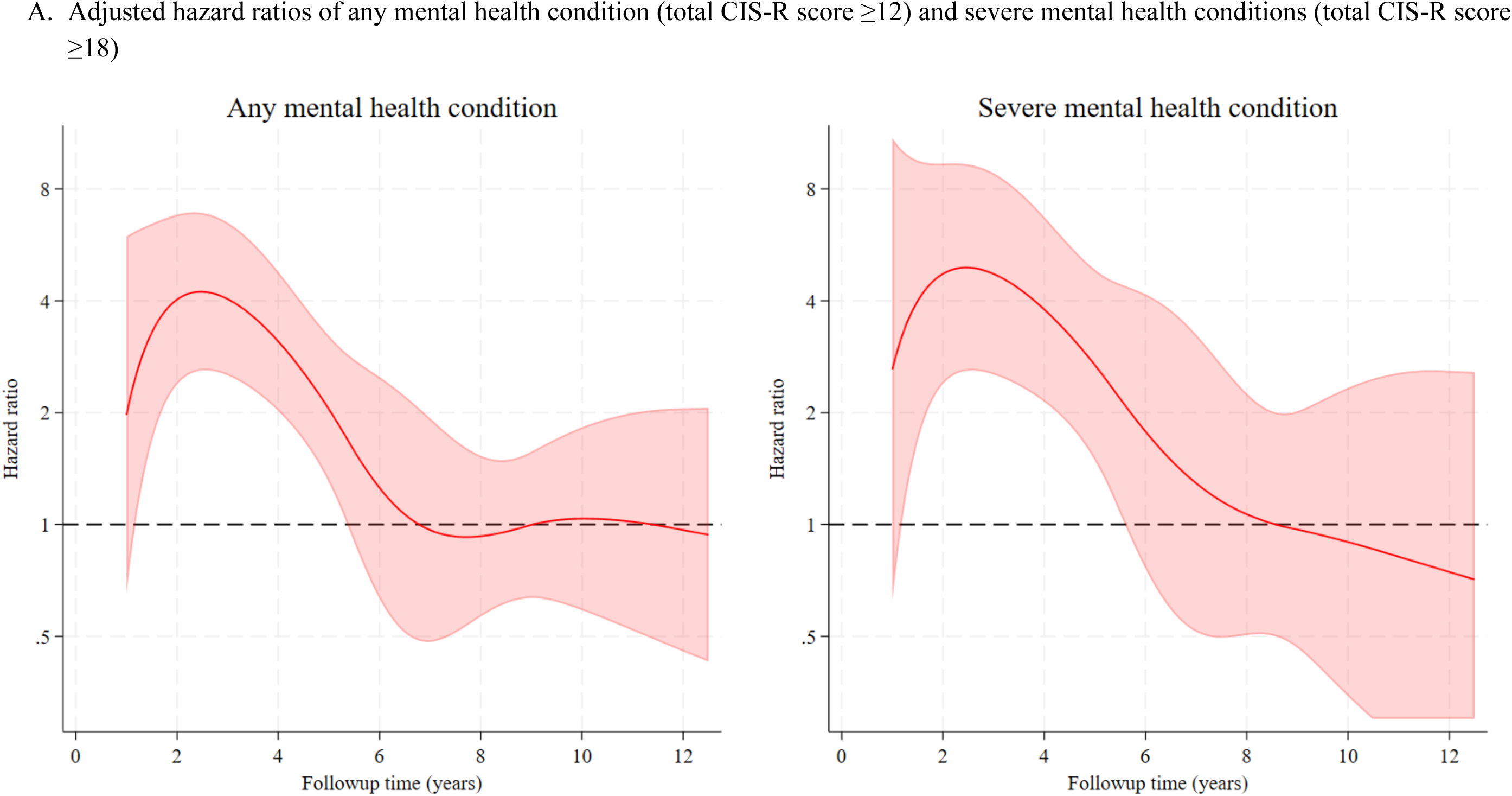
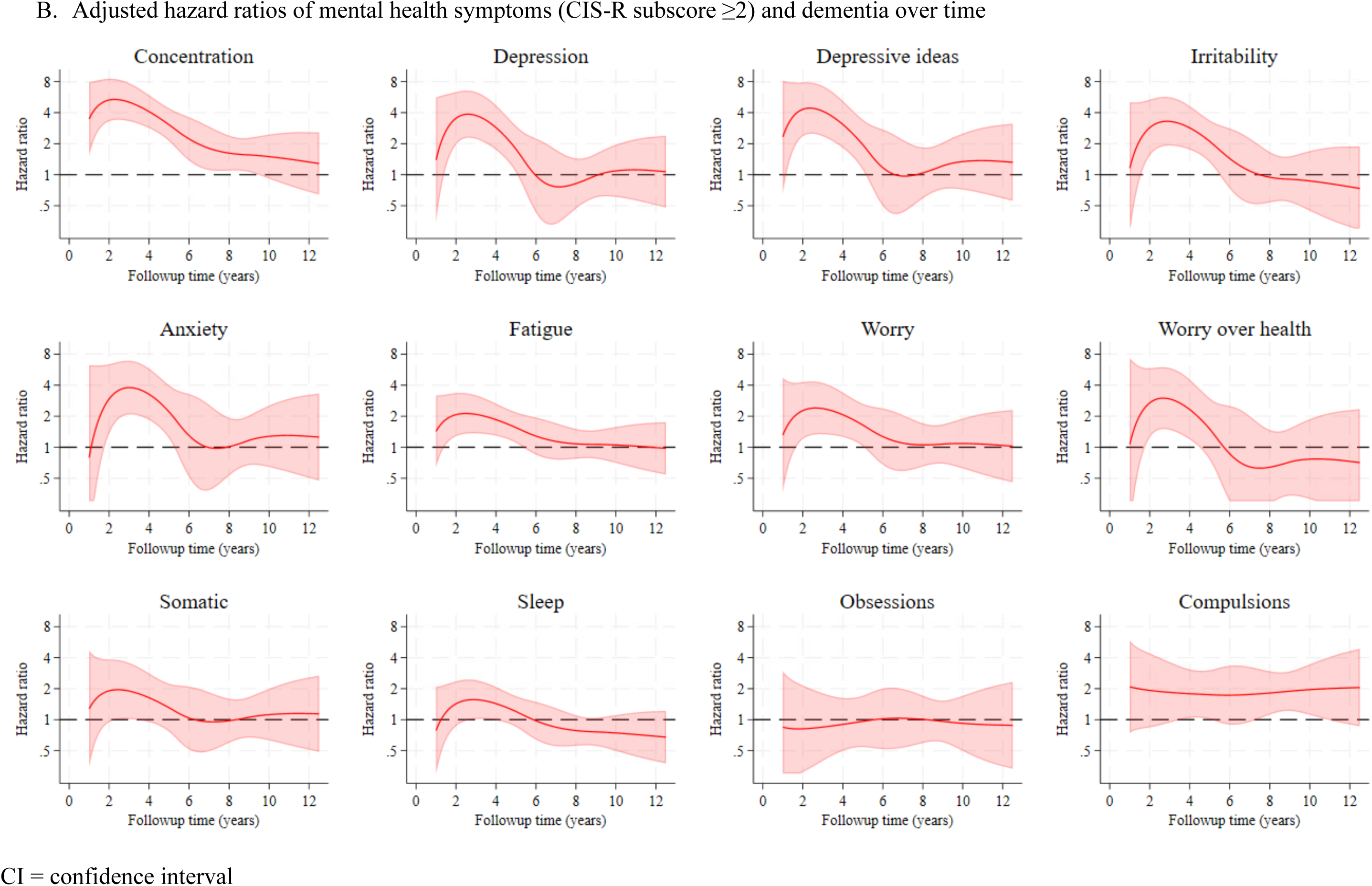
Adjusted hazard ratios of dementia by mental health symptom over 12-year follow-up period Hazard ratios and their confidence intervals were estimated from flexible parametric models adjusted for age, sex, ethnicity, marital status, education, fruit and vegetable intake, physical activity, alcohol consumption, smoking, BMI, diabetes, hypertension, and multimorbidity Notes: Shaded area indicates 95% CI. First year of follow-up was truncated due to few events. If lower confidence intervals fell below 0.3 they were truncated at 0.3.

The average hazard ratio over time for the association between any mental health condition and dementia was 1.63 (95% CI: 1.22, 2.17). The hazard ratio for dementia associated with any mental health condition increased in the 3 years after the CIS-R measure, then decreased to reach no association 6 or more years after (Figure 2A). The hazard ratio at 3 years of follow-up was 4.04 (95% CI: 2.53, 6.50) and at 6 years was 1.26 (95% CI: 0.63, 2.49); estimates at 3 years are provided in Table 3 and for other years of follow-up in Supplemental Table 3. This pattern of association over time was also observed for severe mental health conditions (HR at 3 years: 4.73; 95% CI: 2.54, 8.81); Table 3 and Figure 2A) and for concentration problems, depression, depressive ideas, irritability, anxiety, fatigue, worry, and worry over physical health (Figure 2B). There was no evidence of an association at any time for the following symptoms: somatic symptoms, sleep problems, obsessions, or compulsions (Figure 2B). Supplementary Tables 3a and 3b show the hazard ratio for dementia at each year of follow-up associated with each mental health subscore.

### 3.4. Sensitivity analysis

Using Cox regression, the adjusted hazard ratio for dementia associated with any mental health condition was 1.70 (95% CI: 1.24, 2.33) in the IPW analysis, as compared to the unweighted hazard ratio of 1.63 (95% CI: 1.22, 2.17), indicating the results were minimally affected by attrition bias. This was also the case for other mental health symptoms, as shown in Supplemental Table 4.

## 4. DISCUSSION

Using data on 5,495 individuals with follow-up for a median of 11 years, we found that mental health symptoms increased in the years preceding dementia. The overall CIS-R score increased in the 6 years preceding a dementia diagnosis, as did the symptom subscores of concentration problems, fatigue, depression, irritability, depressive ideas, worry over physical health, and anxiety. We also found that any mental health condition, severe mental health conditions, and the symptoms of concentration problems, depression, depressive ideas, irritability, anxiety, fatigue, worry, and worry over physical health were all associated with higher risk of dementia in the short term, but not the long term, suggesting that these are likely preclinical signs of dementia.

The results of this study add to the existing discussion of mental health symptoms as preclinical signs of dementia and clarified which previously understudied specific mental health symptoms may be prodroma of dementia. Two recent studies have assessed a variety of mental health symptoms as preclinical signs of dementia [12,24], using routinely collected health data which is likely to capture more-severe diagnoses for mental health symptoms than the CIS-R. One study using German insurance claims data to assess risk factors and prodromal features of dementia up to 10 years before dementia found that a diagnosis of anxiety, depression, fatigue, or sleeping disorder were all associated with higher odds of dementia in the 1 year, 2-4 years, and 5-10 years preceding a dementia diagnosis [24]. The odds ratios of anxiety and depression were higher with proximity to diagnosis, though they remained stable for fatigue. Another case-control study using electronic health records for individuals in East London similarly assessed several symptoms including depression, anxiety, insomnia, and fatigue, over three time periods (<2 years, 2-<5 years, 5-<10 years before dementia diagnosis) [12]. They found that depression and anxiety were associated with dementia at all time points, and the association was stronger closer to diagnosis, though fatigue was only associated with incident dementia in the 5-10 years before diagnosis. Our study’s findings align with this evidence in that we found that depression and anxiety increased during the 6 years preceding a dementia diagnosis and were each associated with incident dementia in the short term (∼5 years of follow-up). We observed the same trends for fatigue, however, contrasting the above studies’ results.

Other studies have also found that the association between mental health symptoms is stronger closer to dementia diagnosis [5,15,25]. A review of the association between psychiatric symptoms and dementia found shorter follow-up periods has stronger associations compared to those with longer follow-up periods [15]. A UK Biobank study assessed loneliness, depressed mood, and mood swings, and found that many of the factors had higher odds ratios closer to diagnosis of incident dementia [5].

We found the CIS-R concentration subscore to be the most strongly associated with dementia, with an average 0.72 increase in subscore in the 6 years before dementia diagnosis.

We also found a 5-times increased hazard of dementia in the short term among those with concentration problems. Concentration problems are understood to be early symptoms of dementia [26], which is corroborated by the results of the present study. We also found the CIS-R depression subscore to be an indicator of dementia risk in the short term, aligning with other evidence of depression being a preclinical sign of dementia [6,8,27].

Our findings on anxiety increasing prior to dementia, suggests that it may also be preclinical signs of dementia. A review found that anxiety was associated with dementia and the association increased with increasing age, leading the authors to propose that anxiety is a preclinical sign of dementia rather than a risk factor [28]. Other studies corroborate these findings showing associations of anxiety [29] and worry [30] with cognitive decline, and associations between anxiety and incident dementia among those with cognitive impairment [31]. The findings in the present study are consistent with this previous evidence as we found that worry and anxiety were both associated with dementia in the short term but not the long term. Nonetheless, a moderate long-term association between specific depressive and anxiety symptoms in midlife, measured using the General Health Questionnaire in those with depression, and a higher 20-year dementia risk has been observed in the Whitehall II study [14]. Likewise, in the HUNT study, individuals who later developed dementia had a higher prevalence of mixed anxiety-depressive symptoms more than three decades before their diagnosis [32].

We did not find associations between sleep problems, somatic symptoms, obsessions, or compulsions and dementia, suggesting they may not prodroma to dementia, or at least may not be implicated in dementia in the 6 years before diagnosis. The evidence on sleep and dementia is mixed, however, and difficult to compare as studies use different measures of sleep. Some studies provide evidence of an association between short sleep duration with dementia [33,34], and others providing more inconsistent results for sleep disturbances and dementia risk [35]. The inconsistencies in different measures of sleep problems and results for risk with dementia point to the need for more research to clarify this relationship.

There are several mechanisms through which mental health symptoms and dementia may be associated. Firstly, incipient dementia could cause mental health conditions through neurobiological changes during the development of dementia. One review argues that the buildup of Alzheimer’s proteins, neuronal atrophy, and synaptic degeneration cause mental health conditions commonly seen in patients with Alzheimer’s [36]. As the brain becomes less resilient, then people may be more likely to develop or have worsening mental health disorders. Secondly, dementia and mental health conditions could have common causes which increase the likelihood of them co-occurring. Some authors propose that neuropsychiatric signs may be a manifestation of brain vulnerability, which lowers the threshold for neuronal atrophy to lead to dementia [3]. Cerebrovascular dysfunction is also a proposed common cause between depression and dementia [37]. There are several common neurological changes that are seen among patients with mental health conditions and patients with dementia [36]. A final postulated mechanism is that mental health symptoms cause dementia, for example by depression causing hippocampal atrophy [36] or mental health disorder-induced stress and low-grade chronic inflammation causing downstream neurobiological changes leading to neuronal atrophy [28,38]. However, this hypothesis is inconsistent with our findings as mental health problems were significantly associated with increased dementia risk only in the 5 years following mental health measures, a period during which the underlying pathophysiological processes of dementia are already present, though we only had a maximum 12 years of follow-up.

Major strengths of this study are its robust capture of mental health symptoms using a tool validated in this population [39,40] and longitudinal data which allows for the temporal relationship between mental health symptoms and dementia diagnosis to be evaluated. However, there are limitations. There could be misclassification of the exposure if those with preclinical dementia responded to the CIS-R differently than those without preclinical dementia, through memory impairment associated with dementia. Participants with preclinical dementia may underestimate their mental health symptoms, which would bias the results towards the null. We did not have access to linkage to primary care data, and therefore we may have missed participants who received a dementia diagnosis in primary care only, although we know this is relatively rare. Due to lack of accurately documented dementia subtypes in electronic healthcare records, we could not distinguish between dementia types, and it is likely that different dementia types have different psychological manifestations in the preclinical phase. We did not adjust for multiple comparisons because all individual comparisons were pre-planned and hypothesis driven. We had generally small number of participants with each mental health symptom and dementia, leading to wide confidence intervals and some p-values with borderline interpretability. Nevertheless, the consistency from the two analyses in specific mental health symptoms being relevant in the years before dementia diagnosis increase our confidence that these symptoms are implicated in early dementia. Finally, the Whitehall II population is a British sample of those who were working in the civil service in 1985 and who are primarily White males, which limits the generalizability of these results.

## 5. CONCLUSION

The landscape of dementia diagnosis and treatment is changing rapidly, with the recent approvals of blood-based biomarkers and new drugs [41–43], necessitating timely diagnosis and care planning, support, and other pharmacological and non-pharmacological interventions. Awareness of changes in mental health as a potentially early sign of dementia can highlight individuals who may warrant monitoring and further testing, including with emerging blood-based biomarker diagnostic tools [44]. Using mental health symptoms to support earlier dementia diagnosis may benefit the people who are reluctant to disclose cognitive changes to their family or clinicians [45].

Overall, these results indicate that new or worsening mental health symptoms may represent preclinical signs of dementia, and that some specific symptoms are related to higher risk of dementia. When older adults present with mental health symptoms, clinicians should be aware of the possibility of preclinical dementia and be vigilant for cognitive changes. Timely and accurate diagnosis of dementia in clinical settings remains of utmost importance to provide information and support to those affected, including early intervention and treatment, participation in clinical trials, cost saving, and time to make decisions and a coordinated care plan [46,47].

## Data Availability

All data produced in the present work are contained in the manuscript.

## ACKNOWLEDGEMENTS

We thank the Whitehall II participants, research team, and support staff for carrying out the study and granting us access to the data and Dr. Sujit Rathod for his contributions during the inception phase of the study.

Whitehall II data are available to bona fide researchers for research purposes. Please refer to the Whitehall II data sharing policy at http://www.ucl.ac.uk/whitehallII/data-sharing.

## REFERENCES

1. Cummings J, Apostolova L, Rabinovici GD, Atri A, Aisen P, Greenberg S, et al. Lecanemab: Appropriate Use Recommendations. J Prev Alzheimers Dis. 2023;10(3):362–77.

2. Arvanitakis Z, Shah RC, Bennett DA. Diagnosis and Management of Dementia: A Review. JAMA. 2019 Oct 22;322(16):1589–99.

3. Jang JY, Ho JK, Blanken AE, Dutt S, Nation DA. Affective Neuropsychiatric Symptoms as Early Signs of Dementia Risk in Older Adults. J Alzheimer’s Dis. 2020 Sept 29;77(3):1195–207.

4. Ismail Z, Gatchel J, Bateman DR, Barcelos-Ferreira R, Cantillon M, Jaeger J, et al. Affective and emotional dysregulation as pre-dementia risk markers: exploring the mild behavioral impairment symptoms of depression, anxiety, irritability, and euphoria. Int Psychogeriatr. 2018 Feb;30(2):185–96.

5. You J, Guo Y, Wang YJ, Zhang Y, Wang HF, Wang LB, et al. Clinical trajectories preceding incident dementia up to 15 years before diagnosis: a large prospective cohort study. Mol Psychiatry. 2024 Oct;29(10):3097–105.

6. Singh-Manoux A, Dugravot A, Fournier A, Abell J, Ebmeier K, Kivimäki M, et al. Trajectories of Depressive Symptoms Before Diagnosis of Dementia: A 28-Year Follow-up Study. JAMA Psychiatry. 2017 July 1;74(7):712–8.

7. Amieva H, Le Goff M, Millet X, Orgogozo JM, Pérès K, Barberger-Gateau P, et al. Prodromal Alzheimer’s disease: successive emergence of the clinical symptoms. Ann Neurol. 2008 Nov;64(5):492–8.

8. Almeida OP, Hankey GJ, Yeap BB, Golledge J, Flicker L. Depression as a modifiable factor to decrease the risk of dementia. Transl Psychiatry. 2017 May;7(5):e1117–e1117.

9. Kaup AR, Byers AL, Falvey C, Simonsick EM, Satterfield S, Ayonayon HN, et al. Trajectories of Depressive Symptoms in Older Adults and Risk of Dementia. JAMA Psychiatry. 2016 May 1;73(5):525–31.

10. Chen D, Zhu Z, Shi Y, Li H, Lin R. Prevalence and risk factors for depressive symptoms in older adults with preclinical Alzheimer’s disease: A systematic review and meta-analysis. Int J Nurs Stud. 2026 Jan 1;173:105259.

11. Terracciano A, Luchetti M, Karakose S, Miller AA, Stephan Y, Sutin AR. Meta-analyses of personality change from the preclinical to the clinical stages of dementia. Ageing Res Rev. 2025 Dec 1;112:102852.

12. Zabihi S, Bestwick JP, Jitlal M, Bothongo PL, Zhang Q, Carter C, et al. Early presentations of dementia in a diverse population. Alzheimers Dement. 2025;21(2):e14578.

13. Livingston G, Huntley J, Liu KY, Costafreda SG, Selbæk G, Alladi S, et al. Dementia prevention, intervention, and care: 2024 report of the Lancet standing Commission. The Lancet. 2024 Aug 10;404(10452):572–628.

14. Frank P, Singh-Manoux A, Pentti J, Batty GD, Sommerlad A, Steptoe A, et al. Specific midlife depressive symptoms and long-term dementia risk: a 23-year UK prospective cohort study. Lancet Psychiatry [Internet]. 2025 Dec 15 [cited 2026 Jan 3];0(0). Available from: https://www.thelancet.com/journals/lanpsy/article/PIIS2215-0366(25)00331-1/fulltext

15. Stafford J, Chung WT, Sommerlad A, Kirkbride JB, Howard R. Psychiatric disorders and risk of subsequent dementia: Systematic review and meta-analysis of longitudinal studies. Int J Geriatr Psychiatry. 2022 May;37(5).

16. Butler LM, Houghton R, Abraham A, Vassilaki M, Durán-Pacheco G. Comorbidity Trajectories Associated With Alzheimer’s Disease: A Matched Case-Control Study in a United States Claims Database. Front Neurosci [Internet]. 2021 Oct 8 [cited 2025 May 12];15. Available from: https://www.frontiersin.org/journals/neuroscience/articles/10.3389/fnins.2021.749305/full

17. Hendriks S, Peetoom K, Tange H, van Bokhoven MA, van der Flier WM, Bakker C, et al. Pre-Diagnostic Symptoms of Young-Onset Dementia in the General Practice up to Five Years Before Diagnosis. J Alzheimers Dis JAD. 2022;88(1):229–39.

18. Lewis G, Pelosi AJ, Araya R, Dunn G. Measuring psychiatric disorder in the community: a standardized assessment for use by lay interviewers. Psychol Med. 1992 May;22(2):465–86.

19. Head J, Stansfeld SA, Ebmeier KP, Geddes JR, Allan CL, Lewis G, et al. Use of self-administered instruments to assess psychiatric disorders in older people: validity of the General Health Questionnaire, the Center for Epidemiologic Studies Depression Scale and the self-completion version of the revised Clinical Interview Schedule. Psychol Med. 2013 Dec;43(12):2649–56.

20. Sommerlad A, Perera G, Singh-Manoux A, Lewis G, Stewart R, Livingston G. Accuracy of general hospital dementia diagnoses in England: Sensitivity, specificity, and predictors of diagnostic accuracy 2008–2016. Alzheimers Dement. 2018 July 1;14(7):933–43.

21. Sommerlad A, Perera G, Singh-Manoux A, Lewis G, Stewart R, Livingston G. Accuracy of general hospital dementia diagnoses in England: Sensitivity, specificity, and predictors of diagnostic accuracy 2008–2016. Alzheimers Dement. 2018 July;14(7):933–43.

22. Oberg AL, Mahoney DW. Linear Mixed Effects Models. In: Ambrosius WT, editor. Topics in Biostatistics [Internet]. Totowa, NJ: Humana Press; 2007 [cited 2025 May 12]. p. 213–34. Available from: 10.1007/978-1-59745-530-5_11

23. Muggeo VMR. Estimating regression models with unknown break-points. Stat Med. 2003;22(19):3055–71.

24. Teipel S, Akmatov M, Michalowsky B, Riedel-Heller S, Bohlken J, Holstiege J. Timing of risk factors, prodromal features, and comorbidities of dementia from a large health claims case–control study. Alzheimers Res Ther. 2025 Jan 16;17(1):22.

25. Tapiainen V, Hartikainen S, Taipale H, Tiihonen J, Tolppanen AM. Hospital-treated mental and behavioral disorders and risk of Alzheimer’s disease: A nationwide nested case-control study. Eur Psychiatry J Assoc Eur Psychiatr. 2017 June;43:92–8.

26. Symptoms of dementia [Internet]. nhs.uk. 2023 [cited 2025 Aug 27]. Available from: https://www.nhs.uk/conditions/dementia/symptoms-and-diagnosis/symptoms/

27. Zabihi S, Jeraldo RIE, Anjum R, Carter C, Roche M, Bestwick JP, et al. Pre-diagnostic manifestations of Alzheimer’s disease: A systematic review and meta-analysis of longitudinal studies. Alzheimers Dement. 2025 Jan 3;20(Suppl 3):e085949.

28. Gulpers B, Ramakers I, Hamel R, Köhler S, Oude Voshaar R, Verhey F. Anxiety as a Predictor for Cognitive Decline and Dementia: A Systematic Review and Meta-Analysis. Am J Geriatr Psychiatry. 2016 Oct 1;24(10):823–42.

29. Fung AWT, Lee JSW, Lee ATC, Lam LCW. Anxiety symptoms predicted decline in episodic memory in cognitively healthy older adults: A 3-year prospective study. Int J Geriatr Psychiatry. 2018;33(5):748–54.

30. Pietrzak RH, Maruff P, Woodward M, Fredrickson J, Fredrickson A, Krystal JH, et al. Mild Worry Symptoms Predict Decline in Learning and Memory in Healthy Older Adults: A 2-Year Prospective Cohort Study. Am J Geriatr Psychiatry. 2012 Mar 1;20(3):266–75.

31. Rosenberg PB, Mielke MM, Appleby BS, Oh ES, Geda YE, Lyketsos CG. The Association of Neuropsychiatric Symptoms in MCI with Incident Dementia and Alzheimer Disease. Am J Geriatr Psychiatry. 2013 July 1;21(7):685–95.

32. Aunsmo RH, Strand BH, Anstey KJ, Bergh S, Kivimäki M, Köhler S, et al. Associations between depression and anxiety in midlife and dementia more than 30 years later: The HUNT Study. Alzheimers Dement Amst Neth. 2024;16(4):e70036.

33. Sabia S, Fayosse A, Dumurgier J, van Hees VT, Paquet C, Sommerlad A, et al. Association of sleep duration in middle and old age with incidence of dementia. Nat Commun. 2021 Apr 20;12(1):2289.

34. Šagud M, Bajs Janović M, Uzun S, Kosanović Rajačić B, Kozumplik O, Pivac N. Could self-reporting sleep duration become an important tool in the prediction of dementia? Expert Rev Neurother. 2025 July 3;25(7):737–51.

35. Wong R, Lovier MA. Sleep Disturbances and Dementia Risk in Older Adults: Findings From 10 Years of National U.S. Prospective Data. Am J Prev Med. 2023 June 1;64(6):781–7.

36. Van Dam D, Vermeiren Y, Dekker AD, Naudé PJW, De Deyn PP. Neuropsychiatric Disturbances in Alzheimer’s Disease: What Have We Learned from Neuropathological Studies? Curr Alzheimer Res. 2016 Oct;13(10):1145–64.

37. Geda YE, Schneider LS, Gitlin LN, Miller DS, Smith GS, Bell J, et al. Neuropsychiatric symptoms in Alzheimer’s disease: past progress and anticipation of the future. Alzheimers Dement J Alzheimers Assoc. 2013 Sept;9(5):602–8.

38. Ismail Z, Gatchel J, Bateman DR, Barcelos-Ferreira R, Cantillon M, Jaeger J, et al. Affective and emotional dysregulation as pre-dementia risk markers: exploring the mild behavioral impairment symptoms of depression, anxiety, irritability, and euphoria. Int Psychogeriatr. 2018 Feb;30(2):185–96.

39. Das-Munshi J, Castro-Costa E, Dewey ME, Nazroo J, Prince M. Cross-cultural factorial validation of the Clinical Interview Schedule – Revised (CIS-R); findings from a nationally representative survey (EMPIRIC). Int J Methods Psychiatr Res. 2014;23(2):229–44.

40. Head J, Stansfeld SA, Ebmeier KP, Geddes JR, Allan CL, Lewis G, et al. Use of self-administered instruments to assess psychiatric disorders in older people: validity of the General Health Questionnaire, the Center for Epidemiologic Studies Depression Scale and the self-completion version of the revised Clinical Interview Schedule. Psychol Med. 2013 Dec;43(12):2649–56.

41. What is donanemab? | Alzheimer’s Society [Internet]. [cited 2025 Nov 11]. Available from: https://www.alzheimers.org.uk/blog/what-is-donanemab-alzheimers-drug

42. What is lecanemab? | Alzheimer’s Society [Internet]. [cited 2025 Nov 11]. Available from: https://www.alzheimers.org.uk/blog/what-lecanemab

43. Researching new drugs for Alzheimer’s disease | Alzheimer’s Society [Internet]. [cited 2025 Nov 11]. Available from: https://www.alzheimers.org.uk/what-we-do/researchers/news/researching-new-drugs-alzheimers-disease

44. Frisoni GB, Hansson O, Nichols E, Garibotto V, Schindler SE, Flier WM van der, et al. New landscape of the diagnosis of Alzheimer’s disease. The Lancet. 2025 Sept 27;406(10510):1389–407.

45. Parker M, Barlow S, Hoe J, Aitken L. Persistent barriers and facilitators to seeking help for a dementia diagnosis: a systematic review of 30 years of the perspectives of carers and people with dementia. Int Psychogeriatr. 2020 May;32(5):611–34.

46. Dubois B, Padovani A, Scheltens P, Rossi A, Dell’Agnello G. Timely Diagnosis for Alzheimer’s Disease: A Literature Review on Benefits and Challenges. J Alzheimers Dis JAD. 2016;49(3):617–31.

47. Couch E, Co M, Albertyn CP, Prina M, Lawrence V. A qualitative study of informal caregiver perceptions of the benefits of an early dementia diagnosis. BMC Health Serv Res. 2024 Apr 24;24(1):508.

48. Abe IM, Goulart AC, Santos WR, Lotufo PA, Benseñor IM. Validation of a stroke symptom questionnaire for epidemiological surveys. São Paulo Med J. 2010 July 1;128(4):225–31.

