## Supplemental Materials for "Mental health symptoms as preclinical indicators of dementia: a Whitehall II cohort study"

**SUPPLEMENT**

Supplementary methods

Description of inverse probability weighting procedure

We firstly used logistic regression to estimate the probability of being in the sample (i.e. of having completed the CIS-R in 2012-13), using the following demographic, lifestyle, and health-related variables: age, ethnicity, sex, marital status, education, fruit and vegetable intake, physical activity, diabetes, hypertension, multimorbidity, depression, other mental health disorder, and death during follow-up. There were a few participants missing data on the above variables, so we did a single imputation to impute the missing values. We assessed various interaction terms between demographic variables and all other variables by stepwise selection (alpha <0.05) to create a model that best predicted being in the sample. The inverse of the estimated probabilities was used as weights in the adjusted Cox regression model.

Table 1. Definition of clinical conditions ascertained from linked records

| Condition | Hospital Episode Statistics data ICD-10 (-9) codes [exclusions] | Whitehall II Records (results from previous phases) | Mental Health Services Dataset | Mortality Records | Cancer Registry |
|---|---|---|---|---|---|
| Cancer | CXX [C44] | | | | Same ICD codes |
| Chronic kidney disease | N18 (585) | | | | |
| Chronic obstructive pulmonary disease | J41, J42, J43, J44 (491, 492, 496) | | | | |
| Coronary heart disease | I20, I21, I22, I23, I24, I25 (410, 411, 412, 413, 414) | 12 lead resting electrocardiogram recording | | Same ICD codes | |
| Dementia | F00-03, F05.1, G30-31 | | Same ICD codes | Same ICD codes | |
| Diabetes | E11 (250) | Report by doctor, use of diabetic drugs,<br>fasting glucose ≥7mmol/L, longstanding illness questionnaire | | | |
| Heart failure | I50 (428) | | | | |
| Hypertension | I10, I11, I12, I13, I15 (401, 402, 403, 404, 405) | Blood pressure measures, use of antihypertensive drugs | | | |
| Liver disease | K7X (571) | | | | |
| Parkinson's | G20 (332) | | | Same ICD codes | |
| Rheumatoid arthritis | M05, M06 | | | | |
| Stroke | I60, I61, I63, I64 (430, 431, 434, 436) | MONICA Ausburg stroke questionnaire [48] (Phases 1-9) | | Same ICD codes | |
| Osteoarthritis | M15, M16, M17, M18, M19 (715) | | | | |

ICD = International Classification of Diseases

Figure 1. Study population flow chart

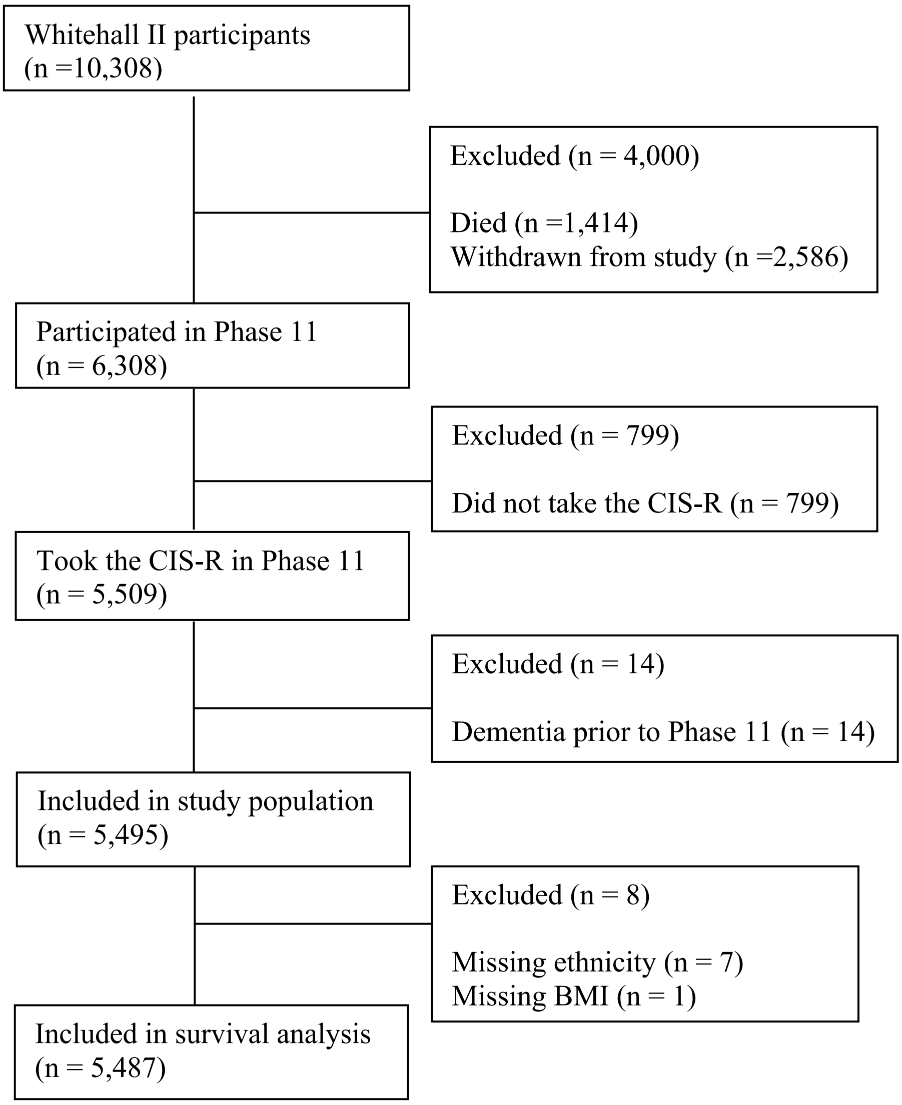


BMI= Body mass index; CIS-R=Clinical Interview Schedule – Revised

Table 2. Prevalence of mental health symptoms (overall and each subscore) at 2015-16, in the full study population and by dementia status at the end of follow-up

| Characteristics | Overall<br>N = 4,609 | No dementia<br>N=4,395 | Dementia<br>N = 314 |
|---|---|---|---|
| Follow-up time (in years), median (IQR) | 8.03 (7.52, 8.54) | 8.09 (7.59, 8.57) | 4.87 (2.27, 6.36) |
| Median total CIS-R score (IQR) | 2 (0, 5) | 2 (0, 4) | 3 (1, 6) |
| Any mental health condition (total CIS-R score ≥12) | 372 (8,1%) | 334 (7.8%) | 38 (12.1%) |
| Severe mental health condition (total CIS-R score ≥18) | 169 (3.7%) | 149 (3.5%) | 20 (6.4%) |
| Symptoms (CIS-R subscore ≥2) | | | |
| Sleep problem symptoms | 1,015 (22.0%) | 960 (21.8%) | 55 (17.5%) |
| Fatigue symptoms | 866 (18.8%) | 785 (18.3%) | 81 (25.8%) |
| Irritability symptoms | 335 (7.3%) | 301 (7.0%) | 34 (10.8%) |
| Worry symptoms | 389 (8.4%) | 358 (8.3%) | 31 (9.9%) |
| Concentration symptoms | 366 (7.9%) | 298 (6.9%) | 68 (21.7%) |
| Obsessions symptoms | 265 (5.7%) | 245 (5.7%) | 20 (6.4%) |
| Depression symptom | 265 (5.7%) | 230 (5.4%) | 35 (11.1%) |
| Somatic symptoms | 277 (6.0%) | 250 (5.8%) | 27 (8.6%) |
| Depressive ideas symptoms | 208 (4.5%) | 183 (4.3%) | 25 (8.0%) |
| Anxiety symptoms | 174 (3.8%) | 157 (3.7%) | 17 (5.4%) |
| Compulsive behaviour symptoms | 160 (3.5%) | 138 (3.2%) | 22 (7.0%) |
| Worry over physical health symptoms | 180 (3.9%) | 161 (3.7%) | 19 (6.1%) |
| Phobia symptoms | 60 (1.3%) | 53 (1.2%) | 7 (2.2%) |
| Panic symptoms | 36 (0.8%) | 34 (0.8%) | 2 (0.6%) |

CIS-R = Clinical Interview Schedule – Revised; IQR = Interquartile Range

Table 3a. Association between any mental health condition (total CIS-R score ≥ 12) and severe mental health condition (total CIS-R score ≥18) and dementia risk at each year of follow-up

| Year of follow-up | Risk of dementia[a] | Any mental health condition Adjusted HR (95% CI) | Severe mental health condition Adjusted HR (95% CI) |
|---|---|---|---|
| 1 | 13/5,495=0.24% | 1.91 (0.65, 5.96) | 2.62 (0.63, 11.0) |
| 2 | 23/5,458=0.42% | 4.03 (2.38, 6.82) | 4.73 (2.39, 9.36) |
| 3 | 21/5,393=0.39% | 4.04 (2.52, 6.50) | 4.73 (2.54, 8.81) |
| 4 | 37/5,316=0.70% | 3.11 (2.02, 4.79) | 3.79 (2.14, 6.72) |
| 5 | 42/5,212=0.81% | 1.04 (1.30, 3.22) | 2.69 (1.49, 4.85) |
| 6 | 47/5,104=0.92% | 1.26 (0.63, 2.49) | 1.78 (0.76, 4.16) |
| 7 | 37/4,991=0.74% | 0.97 (0.48, 1.93) | 1.29 (0.52, 3.24) |
| 8 | 44/4,897=0.90% | 0.93 (0.57, 1.53) | 1.07 (0.50, 2.25) |
| 9 | 49/4,778=1.03% | 1.00 (0.63, 1.57) | 0.97 (0.47, 2.02) |
| 10 | 58/4,642=1.25% | 1.04 (0.59, 1.83) | 0.90 (0.35, 2.33) |
| 11 | 62/4,481=1.38% | 1.02 (0.52, 2.00) | 0.82 (0.26, 2.56) |
| 12.1 | 22/3,703=0.59% | 0.97 (0.46, 2.05) | 0.75 (0.22, 2.59) |

CI = confidence interval; HR = hazard ratio

[a]Numerator: Number of participants with dementia between Y-1 and Y; Denominator: Number of participants at risk for dementia at Y-1

Hazard ratios and their confidence intervals were estimated using flexible parametric models adjusted for age, sex, ethnicity, marital status, education, fruit and vegetable intake, physical activity, alcohol consumption, smoking, BMI, diabetes, hypertension, and multimorbidity.

Table 3b. Association between mental health symptoms (CIS-R subscore ≥ 2) and dementia risk at each year of follow-up

| Year of follow-up | Risk of dementia[a] | Somatic Adjusted HR (95% CI) | Fatigue Adjusted HR (95% CI) | Concentration problems Adjusted HR (95% CI) | Sleep problems Adjusted HR (95% CI) |
|---|---|---|---|---|---|
| 1 | 13/5,495=0.24% | 1.28 (0.35, 4.67) | 2.43 (0.64, 3.19) | 3.49 (1.54, 7.90) | 0.79 (0.30, 2.09) |
| 2 | 23/5,458=0.42% | 1.91 (0.95, 3.85) | 2.07 (1.28, 3.35) | 5.31 (3.31, 8.53) | 1.43 (0.87, 2.36) |
| 3 | 21/5,393=0.39% | 1.90 (1.01, 3.55) | 2.09 (1.37, 3.18) | 5.06 (3.31, 7.73) | 1.57 (1.02, 2.42) |
| 4 | 37/5,316=0.70% | 1.63 (0.95, 2.81) | 1.85 (1.30, 2.65) | 4.10 (2.84, 5.91) | 1.43 (0.99, 2.08) |
| 5 | 42/5,212=0.81% | 1.31 (0.76, 2.24) | 1.55 (1.10, 2.18) | 3.05 (2.12, 4.41) | 1.20 (0.85, 1.69) |
| 6 | 47/5,104=0.92% | 1.04 (0.52, 2.09) | 1.28 (0.85, 1.94) | 2.23 (1.37, 3.61) | 0.98 (0.64, 1.49) |
| 7 | 37/4,991=0.74% | 0.96 (0.50, 1.84) | 1.14 (0.76, 1.71) | 1.81 (1.11, 2.93) | 0.85 (0.56, 1.29) |
| 8 | 44/4,897=0.90% | 0.98 (0.60, 1.60) | 1.08 (0.77, 1.50) | 1.62 (1.10, 2.40) | 0.79 (0.56, 1.11) |
| 9 | 49/4,778=1.03% | 1.06 (0.67, 1.66) | 1.07 (0.78, 1.46) | 1.56 (1.06, 2.28) | 0.76 (0.56, 1.04) |
| 10 | 58/4,642=1.25% | 1.21 (0.63, 1.99) | 1.05 (0.70, 1.57) | 1.50 (0.92, 2.45) | 0.74 (0.50, 1.11) |
| 11 | 62/4,481=1.38% | 1.15 (0.57, 2.32) | 1.03 (0.63, 1.68) | 1.42 (0.78, 2.57) | 0.72 (0.44, 1.18) |
| 12.1 | 22/3,703=0.59% | 1.15 (0.51, 2.57) | 0.99 (0.57, 1.73) | 1.33 (0.68, 2.58) | 0.69 (0.40, 1.21) |

CI = confidence interval; HR = hazard ratio

[a]Numerator: Number of participants with dementia between Y-1 and Y; Denominator: Number of participants at risk for dementia at Y-1

Hazard ratios and their confidence intervals were estimated using flexible parametric models adjusted for age, sex, ethnicity, marital status, education, fruit and vegetable intake, physical activity, alcohol consumption, smoking, BMI, diabetes, hypertension, and multimorbidity.

Table 3b (continued). Association between mental health symptoms (CIS-R subscore ≥ 2) and dementia risk at each year of follow-up

| Year of follow-up | Risk of dementia[a] | Irritability Adjusted HR (95% CI) | Worry over physical health Adjusted HR (95% CI) | Depressive ideas Adjusted HR (95% CI) | Worry Adjusted HR (95% CI) |
|---|---|---|---|---|---|
| 1 | 13/5,495=0.24% | 1.16 (0.27, 5.07) | 1.07 (0.16, 7.32) | 2.33 (0.66, 8.20) | 1.31 (0.37, 4.71) |
| 2 | 23/5,458=0.42% | 2.87 (1.54, 5.35) | 2.70 (1.25, 5.84) | 4.32 (2.37, 7.88) | 2.27 (1.20, 4.29) |
| 3 | 21/5,393=0.39% | 3.29 (1.92, 5.65) | 2.94 (1.48, 5.86) | 4.13 (2.36, 7.23) | 2.37 (1.34, 4.20) |
| 4 | 37/5,316=0.70% | 2.85 (1.75, 4.63) | 2.32 (1.19, 4.53) | 3.08 (1.82, 5.19) | 2.07 (1.26, 3.39) |
| 5 | 42/5,212=0.81% | 2.11 (1.32, 3.37) | 1.50 (0.75, 2.98) | 1.95 (1.12, 3.42) | 1.64 (1.02, 2.64) |
| 6 | 47/5,104=0.92% | 1.44 (0.74, 2.81) | 0.86 (0.30, 2.51) | 1.17 (0.50, 2.75) | 1.27 (0.68, 2.38) |
| 7 | 37/4,991=0.74% | 1.09 (0.53, 2.25) | 0.65 (0.21, 2.00) | 0.97 (0.43, 2.18) | 1.10 (0.59, 2.05) |
| 8 | 44/4,897=0.90% | 0.95 (0.54, 1.66) | 0.64 (0.28, 1.47) | 1.04 (0.59, 1.84) | 1.05 (0.65, 1.71) |
| 9 | 49/4,778=1.03% | 0.91 (0.54, 1.51) | 0.72 (0.34, 1.52) | 1.22 (0.73, 2.05) | 1.07 (0.69, 1.67) |
| 10 | 58/4,642=1.25% | 0.87 (0.45, 1.70) | 0.77 (0.31, 1.87) | 1.34 (0.72, 2.51) | 1.09 (0.62, 1.90) |
| 11 | 62/4,481=1.38% | 0.82 (0.37, 1.84) | 0.76 (0.27, 2.14) | 1.37 (0.66, 2.86) | 1.07 (0.54, 2.11) |
| 12.1 | 22/3,703=0.59% | 0.76 (0.31, 1.88) | 0.73 (0.23, 2.29) | 1.34 (0.59, 3.06) | 1.04 (0.48, 2.25) |

CI = confidence interval; HR = hazard ratio

[a]Numerator: Number of participants with dementia between Y-1 and Y; Denominator: Number of participants at risk for dementia at Y-1

Hazard ratios and their confidence intervals were estimated using flexible parametric models adjusted for age, sex, ethnicity, marital status, education, fruit and vegetable intake, physical activity, alcohol consumption, smoking, BMI, diabetes, hypertension, and multimorbidity.

Table 3b (continued). Association between mental health symptoms (CIS-R subscore ≥ 2) and dementia risk at each year of follow-up

| Year of follow-up | Risk of dementia[a] | Anxiety Adjusted HR (95% CI) | Compulsions Adjusted HR (95% CI) | Obsessions Adjusted HR (95% CI) | Depression Adjusted HR (95% CI) |
|---|---|---|---|---|---|
| 1 | 13/5,495=0.24% | 0.79 (0.10, 6.25) | 2.09 (0.74, 5.88) | 0.85 (0.24, 2.96) | 1.39 (0.35, 5.61) |
| 2 | 23/5,458=0.42% | 2.95 (1.36, 6.39) | 1.93 (0.84, 4.40) | 0.82 (0.31, 2.15) | 3.53 (1.97, 6.35) |
| 3 | 21/5,393=0.39% | 3.79 (2.08, 6.90) | 1.84 (0.94, 3.62) | 0.85 (0.40, 1.80) | 3.76 (2.23, 6.33) |
| 4 | 37/5,316=0.70% | 3.26 (1.84, 5.78) | 1.79 (1.04, 3.08) | 0.90 (0.50, 1.63) | 2.87 (1.75, 4.71) |
| 5 | 42/5,212=0.81% | 2.20 (1.22, 3.99) | 1.75 (1.01, 3.03) | 0.96 (0.54, 1.71) | 1.77 (1.06, 2.97) |
| 6 | 47/5,104=0.92% | 1.30 (0.51, 3.28) | 1.73 (0.90, 3.34) | 1.02 (0.52, 2.01) | 0.98 (0.43, 2.24) |
| 7 | 37/4,991=0.74% | 0.99 (0.38, 2.57) | 1.76 (0.95, 3.25) | 1.03 (0.54, 1.99) | 0.77 (0.34, 1.75) |
| 8 | 44/4,897=0.90% | 1.01 (0.53, 1.93) | 1.82 (1.12, 2.95) | 1.01 (0.59, 1.71) | 0.82 (0.47, 1.44) |
| 9 | 49/4,778=1.03% | 1.17 (0.67, 2.03) | 1.89 (1.22, 2.94) | 0.96 (0.61, 1.53) | 0.98 (0.60, 1.60) |
| 10 | 58/4,642=1.25% | 1.28 (0.64, 2.53) | 1.96 (1.12, 3.44) | 0.92 (0.50, 1.71) | 1.09 (0.61, 1.95) |
| 11 | 62/4,481=1.38% | 1.30 (0.57, 2.96) | 2.01 (0.99, 4.07) | 0.90 (0.41, 1.98) | 1.12 (0.56, 2.21) |
| 12.1 | 22/3,703=0.59% | 1.28 (0.51, 3.22) | 2.04 (0.90, 4.62) | 0.89 (0.35, 2.22) | 1.09 (0.51, 2.35) |

CI = confidence interval; HR = hazard ratio

[a]Numerator: Number of participants with dementia between Y-1 and Y; Denominator: Number of participants at risk for dementia at Y-1

Hazard ratios and their confidence intervals were estimated using flexible parametric models adjusted for age, sex, ethnicity, marital status, education, fruit and vegetable intake, physical activity, alcohol consumption, smoking, BMI, diabetes, hypertension, and multimorbidity.

Table 4. Association between the CIS-R overall mental health condition score and symptom subscores and risk of dementia in original study population and using inverse probability weighted study population

| Condition | Adjusted HR Unweighted (95% CI) | p-value | Adjusted HR Weighted (95% CI) | p-value |
|---|---|---|---|---|
| Any mental health condition (CIS-R subscore ≥12) | 1.63 (1.22, 2.17) | <0.001 | 1.70 (1.24, 2.33) | <0.001 |
| Severe mental health condition (CIS-R subscore ≥18) | 1.91 (1.28, 2.86) | 0.002 | 2.03 (1.32, 3.14) | 0.001 |
| Symptom (CIS-R subscore ≥2) | | | | |
| Somatic | 0.84 (0.56, 1.23) | 0.40 | 1.25 (0.85, 1.83) | 0.256 |
| Fatigue | 1.31 (1.04, 1.66) | 0.021 | 1.28 (1.00, 1.65) | 0.051 |
| Concentration | 2.46 (1.92, 3.14) | <0.001 | 2.46 (1.89, 3.20) | <0.001 |
| Sleep | 0.95 (0.75, 1.19) | 0.60 | 1.02 (0.80, 1.31) | 0.869 |
| Irritability | 1.47 (1.07, 2.03) | 0.017 | 1.53 (1.09, 2.15) | 0.014 |
| Worry over physical health | 1.20 (0.77, 1.87) | 0.40 | 1.27 (0.79, 2.05) | 0.326 |
| Depressive ideas | 1.82 (1.30, 2.57) | <0.001 | 2.02 (1.38, 2.96) | <0.001 |
| Worry | 1.38 (1.01, 1.88) | 0.044 | 1.32 (0.96, 1.83) | 0.090 |
| Anxiety | 2.49 (1.27, 4.87) | 0.008 | 1.77 (1.18, 2.65) | 0.005 |
| Compulsions | 1.90 (1.34, 2.70) | <0.001 | 1.75 (1.19, 2.59) | 0.005 |
| Obsessions | 0.94 (0.65, 1.38) | 0.80 | 0.80 (0.53, 1.22) | 0.305 |
| Depression | 1.51 (1.10, 2.07) | 0.012 | 1.71 (1.21, 2.42) | 0.002 |

HR = Hazard ratio; CI = Confidence interval

Cox regression models adjusted for age, sex, ethnicity, marital status, education, fruit and vegetable intake, physical activity, alcohol consumption, smoking, BMI, diabetes, hypertension, and multimorbidity
